# Pilot Randomized Trial of Intermittent Theta-Burst Stimulation versus H-Coil Transcranial Magnetic Stimulation for Treatment-Resistant Depression

**DOI:** 10.64898/2026.03.13.26348335

**Authors:** Véronique Desbeaumes Jodoin, Emma Bousseau, François Trottier-Duclos, Didier Jutras-Aswad, François Lespérance, Élie Bou Assi, Dang Khoa Nguyen, Daniel M. Blumberger, Travis E. Baker, Martijn Arns, Zafiris J. Daskalakis, Paul Lespérance, Jean-Philippe Miron

## Abstract

**Background:** Intermittent theta burst stimulation (iTBS) and H-coil repetitive transcranial magnetic stimulation (rTMS) are FDA-cleared treatments for major depression; yet their comparative effectiveness in treatment-resistant depression (TRD) has not been evaluated in randomized trials. This pilot randomized trial was designed to obtain preliminary comparative estimates and to explore whether baseline cognitive functioning relates to early remission.

**Methods:** Twenty-eight adults with TRD were randomized to six weeks of iTBS delivered to the dorsolateral prefrontal cortex (DLPFC) using a figure-8 coil (n=15) or H-coil rTMS delivered to the dorsomedial prefrontal cortex (DMPFC) using a H7-coil (n=13). The primary outcome was change in 17-item Hamilton Depression Rating Scale (HRSD-17) score from baseline to week 6, analyzed with ANCOVA. Additional outcomes included response, remission, and symptom trajectories through week 18. Exploratory analyses examined the association between baseline cognitive functioning, such as executive functions and memory, and remission.

**Results:** Twenty-five participants completed all 30 sessions. Adjusted week-6 HRSD-17 scores did not differ between groups (mean difference −0.40, 95% CI −5.23 to 4.43; p=.865). Response rates were 40.0% for iTBS and 50.0% for H-coil (p>.60), and remission rates were identical across groups (20.0%). Remitters showed higher baseline executive functioning than non-remitters in exploratory analyses, although these associations were not confirmed in adjusted models.

**Conclusion:** In this pilot trial, iTBS and H7-coil rTMS showed symptom improvement, with no clear between-group pattern. Exploratory findings suggest a potential signal involving executive functioning that warrants further investigation. These results inform the feasibility and design of larger comparative trials.

**Trial registration:** ClinicalTrials.gov (NCT05902312)

## 1. Introduction

Major depressive disorder (MDD) is a leading cause of disability worldwide, affecting an estimated 300 million people and contributing substantially to morbidity, mortality, and societal costs [1]. Although antidepressants and psychotherapy are effective for many, up to 55% of patients do not achieve remission and meet criteria for treatment-resistant depression (TRD) [2]. Repetitive transcranial magnetic stimulation (rTMS) is an effective treatment for TRD, with pooled response and remission rates of approximately 40–45% and 20–25%, respectively, according to a recent meta-analysis of randomized sham-controlled trials in patients with at least two prior antidepressant treatment failures [3]. According to these estimates, only a minority of patients therefore achieve substantial clinical improvement, reinforcing the need to optimize stimulation parameters.

To that end, new rTMS protocols have been developed. Intermittent theta burst stimulation (iTBS), cleared by the U.S. Food and Drug Administration (FDA) for patients with inadequate response to prior antidepressants since 2018, is now a well-established rTMS protocol [4,5]. Following the large THREE-D trial demonstrating non-inferiority to standard 10-Hz rTMS [6], and given its markedly shorter session duration, iTBS has been increasingly adopted in clinical practice, becoming the most commonly used rTMS protocol for depression in some national settings, such as Sweden [7]. Recent guidelines likewise recognize iTBS as an evidence-based first-line option, equivalent to 10-Hz stimulation [8].

Another rTMS option with demonstrated efficacy involves coil designs that can generate a broader and deeper electromagnetic field than figure-8 coils used for rTMS, such as the H-coils. A pivotal multicenter randomized controlled trial demonstrated the antidepressant efficacy of the H1-coil compared with sham in patients who had not achieved satisfactory improvement from prior antidepressants, leading to FDA clearance in 2013 for patients with inadequate response to prior antidepressants [9]. A subsequent RCT comparing H1-coil rTMS with figure-8 rTMS did find significantly higher response rates and greater reductions in depression severity with H-coil rTMS, while remission rates did not differ significantly between groups [10]. The H7 coil, originally designed for obsessive–compulsive disorder [11], has also demonstrated antidepressant effects and received FDA clearance in 2022 for patients with inadequate response to prior antidepressants, including those with comorbid anxiety symptoms. Compared with both the H1 and figure-8 coils, the H7 generates deeper and broader fields, stimulating an estimated cortical volume of ∼40 cm^3^ versus ∼3 cm^3^ for figure-8 coils [12]. However, this increased stimulated volume comes with a fundamental depth–focality trade-off. Compared with figure-8 coils, H-coils distribute the induced electric field over broader cortical territories, resulting in deeper but less focal stimulation and lower peak electric field strength at any single cortical location. Computational modeling and simulation studies have consistently demonstrated this trade-off across coil designs, highlighting that greater spatial extent is achieved at the expense of focal field intensity [12][13]. Whether broader, distributed stimulation or more focal, higher-intensity stimulation yields superior antidepressant effects remains an empirical question.

Figure-8 coils used for rTMS were initially developed for neurophysiological experimentation, where more focal stimulation is preferred for precise target engagement. Unfortunately, the high individual variability of the dorsolateral prefrontal cortex (DLPFC) and the probabilistic nature of figure-8 coil positioning render them theoretically more prone to targeting error. Neuronavigation systems have been developed to optimize targeting, but these tools are expensive and technically complex, currently precluding widespread use [14]. The H7 coil is primarily thought to engage medial prefrontal regions, particularly the dorsomedial prefrontal cortex DMPFC, with secondary engagement of the DLPFC [15]. Rather than reflecting a simple distinction between superficial and deep stimulation, this line of work highlights a potential contrast between lateral and medial prefrontal targets, which may differentially engage neural circuits relevant to mood regulation. Whether stimulation of medial prefrontal networks confers clinical advantages over lateral targets only remains an open question [16].

In addition to differences in stimulation targets, patient-level factors such as cognitive functioning may also influence neuromodulation outcomes. Many patients with TRD present deficits in executive processes, attention, processing speed and memory, which are linked to poorer functional outcomes and reduced treatment response [17,18]. These cognitive impairments are thought to reflect underlying prefrontal network dysfunction, potentially limiting the brain’s capacity for plastic reorganization in response to stimulation. Accordingly, prefrontal integrity and executive control have been proposed as markers of network plasticity and predictors of response to rTMS-based interventions [19][20]. Although the evidence remains early, baseline cognitive performance could help identify individuals most likely to benefit from neuromodulation.

To our knowledge, no randomized trial has directly compared left-DLPFC iTBS delivered with a figure-8 coil and H7-coil rTMS in TRD. Given the absence of head-to-head data and the pilot design of this study, the goal was to obtain preliminary comparative estimates that could inform the design and power requirements of a future full-scale trial. We therefore examined preliminary clinical outcomes with iTBS and H7-coil rTMS, while also assessing feasibility elements typical of pilot work. As a secondary exploratory aim, we evaluated whether baseline cognitive functioning showed any preliminary association with early remission.

## 2. Methods

### 2.1 Design

This was a pilot, randomized, single-center, two-arm, parallel-group trial conducted at the Centre Hospitalier de l’Université de Montréal (CHUM) between January 2024 and September 2025. The protocol was approved by the institutional review board of the CHUM, and all participants provided written informed consent before enrollment. The study was conducted in accordance with the Declaration of Helsinki and was registered at ClinicalTrials.gov (identifier: NCT05902312). Patients were recruited through medical referrals to the interventional psychiatry clinic at CHUM. Randomization was performed in a 1:1 ratio. As this was a pilot trial, no formal power calculation was undertaken; the sample size was determined to generate preliminary comparative estimates.

### 2.2 Participants

Adults aged 21–70 years with Diagnostic and Statistical Manual of Mental Disorders, 5th Edition (DSM-5) MDD (single or recurrent episode) confirmed by a study psychiatrist were eligible if they had a baseline 17-item Hamilton Depression Rating Scale (HDRS-17) score ≥18 and demonstrated treatment resistance (≥ 2 adequate antidepressant trials per the Antidepressant Treatment History Form [ATHF] score > 3, or documented intolerance to ≥ 2 distinct trials) while on a stable antidepressant regimen for ≥ 4 weeks. Exclusions were prior rTMS exposure; current or past bipolar or psychotic disorder; primary substance use disorder within 3 months (nicotine/cannabis permitted); a primary anxiety or personality disorder judged more impairing than MDD; unstable medical or neurological illness; contraindications to rTMS (e.g., ferromagnetic implants, pacemaker/implanted stimulators); clinically significant laboratory abnormalities; pregnancy; active suicidality requiring urgent intervention; unstable psychotherapy; ongoing anticonvulsant therapy; or benzodiazepine use >2 mg/day lorazepam-equivalent (benzodiazepines permitted only after stimulation sessions). Patients with chronic depression (≥ 2 years) and those with prior electroconvulsive therapy or ketamine exposure were eligible.

### 2.3 Randomization and blinding

Participants were randomized in a 1:1 ratio to receive either iTBS (figure-8) or H7-coil rTMS using a computer-generated allocation list, stratified by level of treatment resistance (≤2 vs >2 failed antidepressant trials). Randomization was generated by an independent researcher not involved in enrollment or assessment. Owing to device characteristics, the trial was open-label for participants and treating clinicians; however, outcome assessors and data analysts remained blinded to treatment allocation.

### 2.4 Intervention

Participants received 30 treatment sessions over six weeks (five sessions per week). In the iTBS rTMS arm, stimulation was delivered over the left dorsolateral prefrontal cortex using a MagPro X100 stimulator (MagVenture) equipped with a B70 fluid-cooled figure-8 coil. The stimulation target was localized using the modified Beam-F3 method. The FDA-cleared iTBS protocol for major depression was applied, consisting of bursts of three pulses at 50 Hz repeated at 5 Hz, for a total of 600 pulses per session, delivered at 120% of the resting motor threshold. In the H7-coil arm, stimulation was delivered using a BrainsWay system equipped with the H7 coil, designed to engage medial prefrontal regions. The FDA-cleared protocol for major depression was applied, consisting of 18 Hz stimulation, 1,980 pulses per session, delivered at 120% of the resting hand motor threshold. Treatment was discontinued if participants missed four or more consecutive sessions, demonstrated a >25% increase in HRSD-17 score at two consecutive visits, developed active suicidality, or at the investigator’s judgment. When feasible, follow-up assessments were pursued even if treatment was discontinued. Adverse events were systematically monitored at each visit using structured queries, and tolerability was assessed throughout the trial.

### 2.5 Outcomes

Feasibility outcomes typical of pilot trials, namely recruitment, retention, and adherence to the treatment schedule, were documented to inform the design of a future full-scale trial. Preliminary clinical outcomes included change in HRSD-17 score from baseline to week 6 (primary clinical outcome). Secondary clinical outcomes consisted of categorical response (≥50% reduction in HRSD-17) and remission (HRSD-17 ≤7) at week 6, as well as depressive symptom trajectories across follow-up assessments using HRSD-28, MADRS, and QIDS-SR-16. Exploratory outcomes examined whether baseline cognitive functioning showed a preliminary association with early remission. Safety outcomes included adverse events, serious adverse events, and treatment discontinuation.

### 2.6 Assessments

#### 2.6.1 Clinical assessments

Assessments were conducted at baseline (week 0), at the end of treatment (week 6), and during follow-up visits at weeks 7, 10, and 18. Depressive symptoms were assessed using the 17-item Hamilton Depression Rating Scale (HRSD-17), the 28-item Hamilton Depression Rating Scale (HRSD-28), the Montgomery–Åsberg Depression Rating Scale (MADRS), and the Quick Inventory of Depressive Symptomatology – Self Report (QIDS-SR-16). Safety was assessed at each visit through systematic monitoring of adverse and serious adverse events.

#### 2.6.2 Cognitive assessments

A brief cognitive battery was administered at baseline to characterize neuropsychological functioning. This included verbal fluency (Controlled Oral Word Association Test), processing speed and cognitive flexibility (Trail Making Test A and B), visuoconstructive abilities and memory (Rey–Osterrieth Complex Figure, copy and immediate recall), and verbal learning and memory (Rey Auditory Verbal Learning Test). A composite executive function score was computed by averaging z-scores of verbal fluency (COWAT), TMT-B, and Rey Complex Figure copy performance, standardized using the baseline sample mean and standard deviation.

### 2.7 Statistical analysis

Feasibility outcomes were summarized descriptively, including recruitment rate, proportion of eligible participants who proceeded to randomization, adherence to the treatment schedule, and retention at post-treatment and follow-up visits.

Given the pilot design and limited sample size, clinical analyses were considered preliminary and were conducted to provide estimates that may inform future full-scale trials rather than to formally test hypotheses of treatment efficacy. The primary clinical outcome, change in HRSD-17 score from baseline to week 6, was examined using an ANCOVA model adjusted for baseline HRSD-17 and treatment-resistance level. Within-group changes over time were also summarized descriptively. Secondary clinical outcomes (response and remission at week 6) and longitudinal depressive symptoms were analyzed to generate preliminary comparative estimates, with group differences interpreted cautiously. Exploratory analyses assessed whether baseline cognitive performance showed a preliminary association with early remission using logistic regression models adjusted for baseline depression severity. Missing data were handled within mixed-effects models under the assumption of missing at random. Effect sizes and 95% confidence intervals are reported to aid interpretation. All analyses are exploratory in keeping with the pilot nature of the study.

## 3. Results

### 3.1 Participants

Of 32 patients screened for eligibility, 29 provided written informed consent (Fig. 1). One patient withdrew before initiating treatment, leaving 28 randomized participants who received at least one treatment session (15 iTBS, 13 H-coil). Three participants discontinued during the treatment course, all in the H-coil group, resulting in 25 completers who finished all 30 sessions (15/15 iTBS; 10/13 H-coil). Treatment adherence was therefore 100% in the iTBS group and 76.9% in the H-coil group, with an overall adherence rate of 89.3% (25/28). Retention at follow-up was also strong: 25 participants completed week-7 and week-10 assessments and 24 providing clinician-rated data at week 18. Baseline demographic and clinical characteristics are summarized in Table 1.

**Figure 1.**
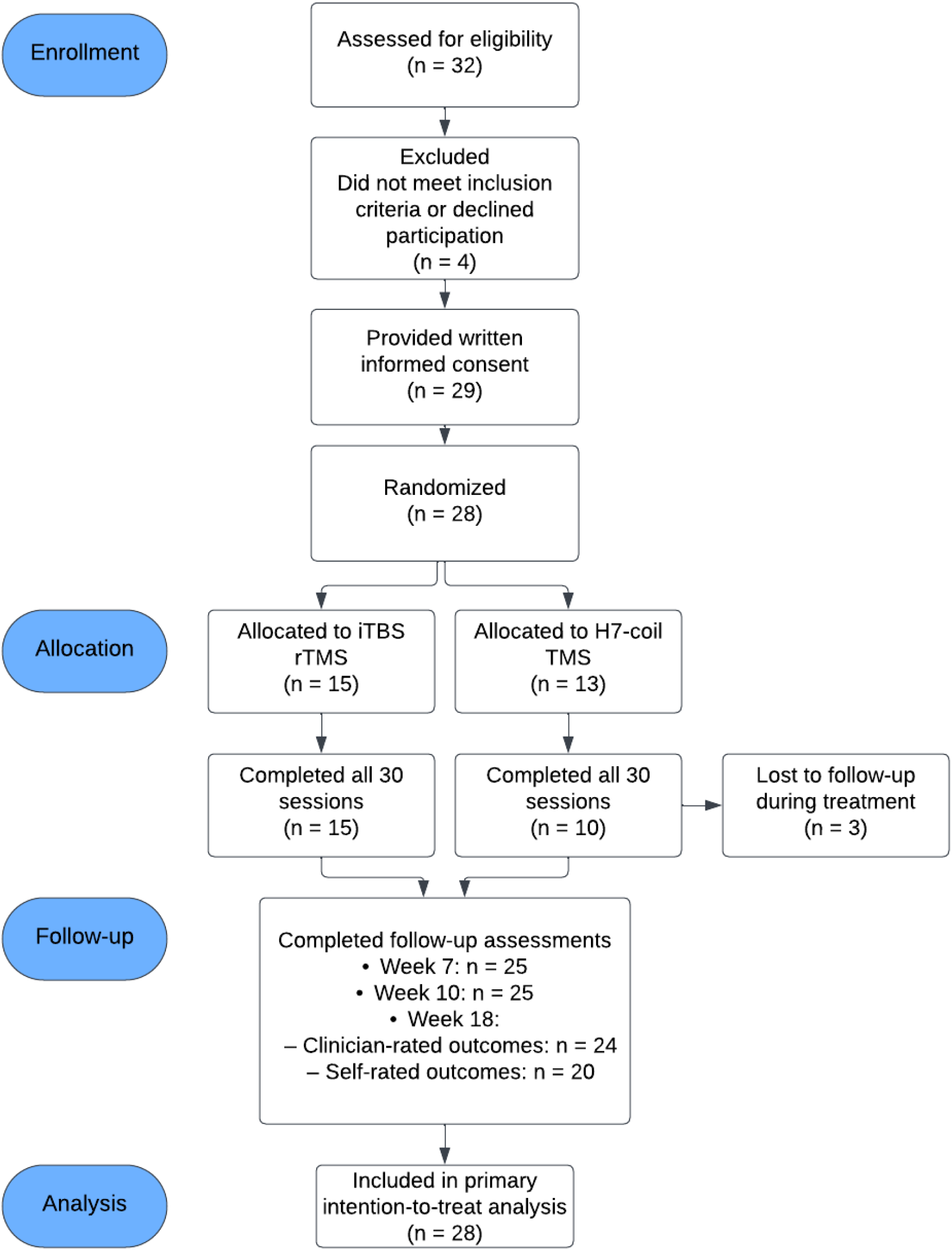
CONSORT flow diagram.

**Table 1.** Baseline demographic and clinical characteristics of participants by treatment group.

| Variables | iTBS (n=15) | dTMS (n=13) | p-value |
| --- | --- | --- | --- |
| **Demographic** |  |  |  |
| Age, years (mean ± SD) | 50.1 ± 11.4 | 45.1 ± 9.4 | .22 |
| Sex, female n (%) | 13 (86.7%) | 10 (76.9%) | .50 |
| **Clinical history** |  |  |  |
| Duration of current episode, months | 39.6 ± 46.2 | 45.8 ± 47.5 | .73 |
| Number of past episodes | 3.1 ± 1.3 | 4.7 ± 3.5 | .10 |
| Resistance (>2 adequate trials) n (%) | 9 (60%) | 6 (46.2%) | .46 |
| **Concomitant medications, n (%)** |  |  |  |
| SSRI | 12 (80%) | 10 (76.9%) | 1.0 |
| SNRI | 9 (60%) | 12 (92.3%) | .08 |
| TCA | 2 (13.3%) | 1 (7.7%) | .63 |
| MAOI | 0 (0%) | 0 (0%) | 1.0 |
| Lithium | 1 (6.7%) | 1 (7.7%) | 1.0 |
| Anticonvulsivant | 1 (6.7%) | 0 (0%) | 1.0 |
| Antipsychotic | 3 (20%) | 3 (23.1%) | .52 |
| **Depression severity** |  |  |  |
| HRSD-17 | 22.1 ± 2.2 | 21.8 ± 3.8 | .80 |
| HRSD-28 | 33.3 ± 4.7 | 31.2 ± 6.0 | .31 |
| MADRS | 28.3 ± 4.7 | 28.2 ± 5.6 | .99 |
| **Cognitive measures** |  |  |  |
| COWAT | 37.7 ± 12.1 | 31.7 ± 7.5 | .13 |
| TMT-A | 25.3 ± 7.3 | 40.3 ± 20.3 | **.01** |
| TMT-B | 67.5 ± 24.3 | 93.7 ± 46.3 | .07 |
| ROCF copy | 34.5 ± 1.5 | 33.6 ± 2.6 | .27 |
| ROCF imm recall | 21.9 ± 6.0 | 20.8 ± 7.2 | .64 |
| RAVLT total learning | 52.2 ± 8.2 | 49.2 ± 12.2 | .45 |
| McNair general cognitive deficits | 68.7 ± 23.4 | 78.8 ± 23.6 | .27 |
| McNair mnesic deficits | 20.2 ± 7.2 | 22.8 ± 8.5 | .40 |

Values are mean ± SD unless otherwise indicated. Between-group comparisons were performed using independent-samples t tests for continuous variables and $\chi^2$ (or Fisher's exact test) for categorical variables.

### 3.2 Primary outcome

At week 6, there was no significant difference between treatment groups in HRSD-17 scores. An ANCOVA controlling for baseline HRSD-17 and level of treatment resistance (ATHF) revealed no main effect of treatment group (F(1,21) = 0.03, p = .865). Adjusted marginal means were 12.3 (95% CI 9.4–15.3) for iTBS and 12.7 (95% CI 9.1–16.4) for H-coil, with an estimated between-group difference of –0.40 points (SE 2.32; 95% CI –5.23 to 4.43).

### 3.3 Secondary outcomes

Analyses of other clinician- and self-rated depression scales (HRSD-28, MADRS, QIDS-SR-16) across all time points produced convergent results, with no significant group differences (all p > .05). At week 6, categorical response was achieved by 40.0% (6/15) of iTBS participants and 50.0% (5/10) of H-coil participants, while remission occurred in 20.0% of each group (3/15 iTBS vs 2/10 H-coil). Group differences were non-significant (χ^2^ and Fisher’s exact tests, all p > .60). Sensitivity analyses counting non-completers as non-responders (ITT) yielded similar proportions for response (40.0% vs 38.5%) and remission (20.0% vs 15.4%), leading to the same qualitative conclusion.

### 3.4 Longitudinal outcomes

Across the follow-up period, there was a significant main effect of time on HRSD-17 scores (F(3,51) = 3.43, p = .024, η^2^p = .17), reflecting overall symptom improvement. Pairwise comparisons indicated a significant reduction between week 6 (M = 13.0, SE = 1.36) and week 10 (M = 9.7, SE = 1.18; p = .028), whereas other contrasts were non-significant after Bonferroni correction. No main effect of group was observed (F(1,17) = 0.63, p = .440, η^2^p = .04), nor a significant time × group interaction (F(3,51) = 0.43, p = .736, η^2^p = .02), indicating that trajectories of improvement did not differ between iTBS and H-coil. Figure 2 illustrates parallel decreases in HRSD-17 scores across both groups, converging toward similar post-treatment levels.

**Figure 1.**
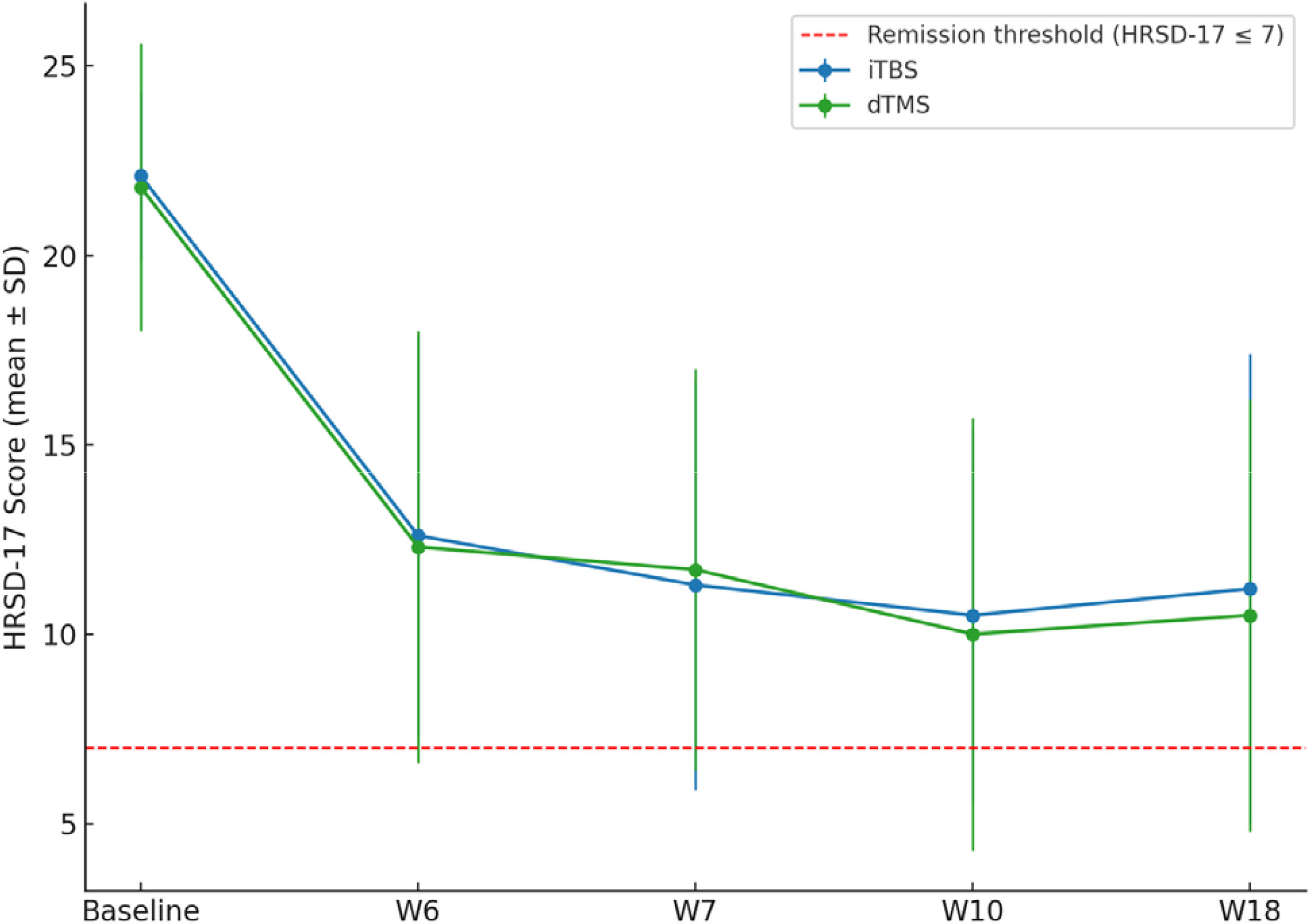
Trajectory of depressive symptoms (HRSD-17 scores) over time in iTBS and dTMS groups. Mean HRSD-17 scores (± standard deviation) are shown at each time point: baseline, week 6 (end of treatment), week 7, week 10, and week 18. The red dashed line indicates the remission threshold (HRSD-17 ≤ 7).

### 3.5 Exploratory cognitive analyses

When considering the whole sample, remitters (MADRS ≤ 10 at week 7) outperformed non-remitters on the executive composite (M = 0.43, SD = 0.73 vs M = –0.16, SD = 0.78; t(23) = –2.12, p = .045, d = 0.69) and showed a trend toward better performance on the COWAT (M = 0.41, SD = 1.08 vs M = –0.23, SD = 0.76; t(23) = –1.69, p = .052, d = 0.94). Stratified analyses suggested that these effects were primarily driven by the iTBS group (executive composite: t(13) = –2.02, p = .065, d = 0.73; COWAT: t(13) = –1.88, p = .083, d = 1.06), whereas no comparable differences were observed within the H-coil TMS group (see Figure 3).

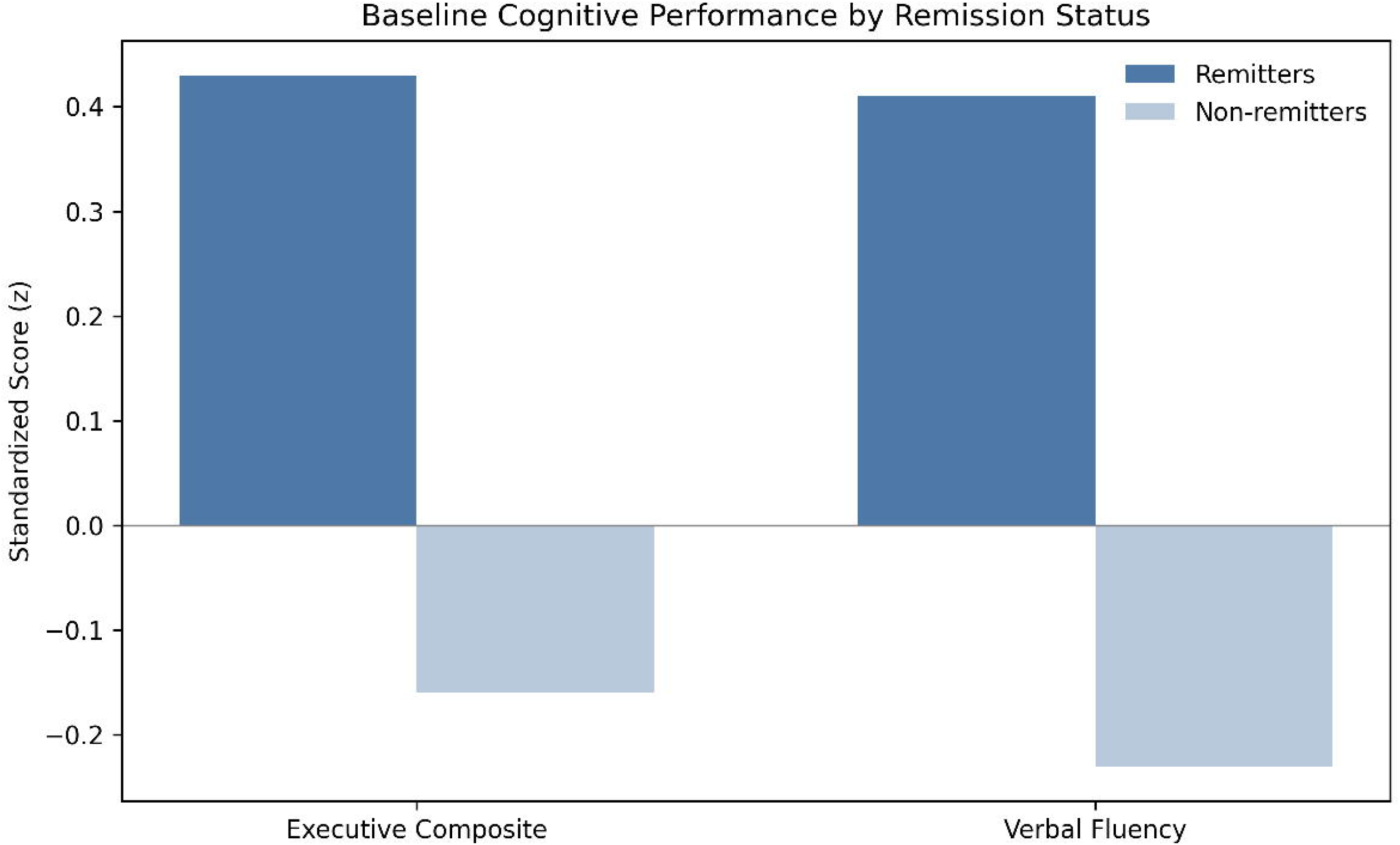

Exploratory logistic regression models including baseline executive functioning, treatment group, their interaction, age, sex, and baseline MADRS score did not reach statistical significance (χ^2^(6) = 6.44, p = .375), although model fit indices suggested moderate explanatory value (Nagelkerke R^2^ = .303; Hosmer–Lemeshow p = .231). None of the individual predictors reached significance, though higher baseline executive functioning showed a positive, non-significant association with remission likelihood (OR = 4.95).

### 3.6 Adverse events

All participants reported at least one adverse event. While stimulation-site discomfort was reported by all participants in both groups (100%), the overall adverse event profile differed between stimulation approaches. Fatigue and headache or migraine were reported by all participants receiving H-coil TMS (100% for both), compared with 46.7% of participants in the iTBS group. Anxiety-related symptoms were reported by 40% of H-coil participants and 33.3% of iTBS participants.

Perceived stimulation-related discomfort was rated as mild to moderate overall, with comparable mean pain ratings between groups (iTBS: 3.91; H-coil TMS: 3.57). No serious adverse events occurred, and no participants discontinued treatment due to adverse effects. Overall, H-coil TMS was associated with a higher frequency of systemic adverse effects, whereas iTBS was better tolerated, despite comparable levels of stimulation-related discomfort (see Figure 4). Given the small number of discontinuations, formal comparisons of adverse event burden between completers and non-completers were not feasible.

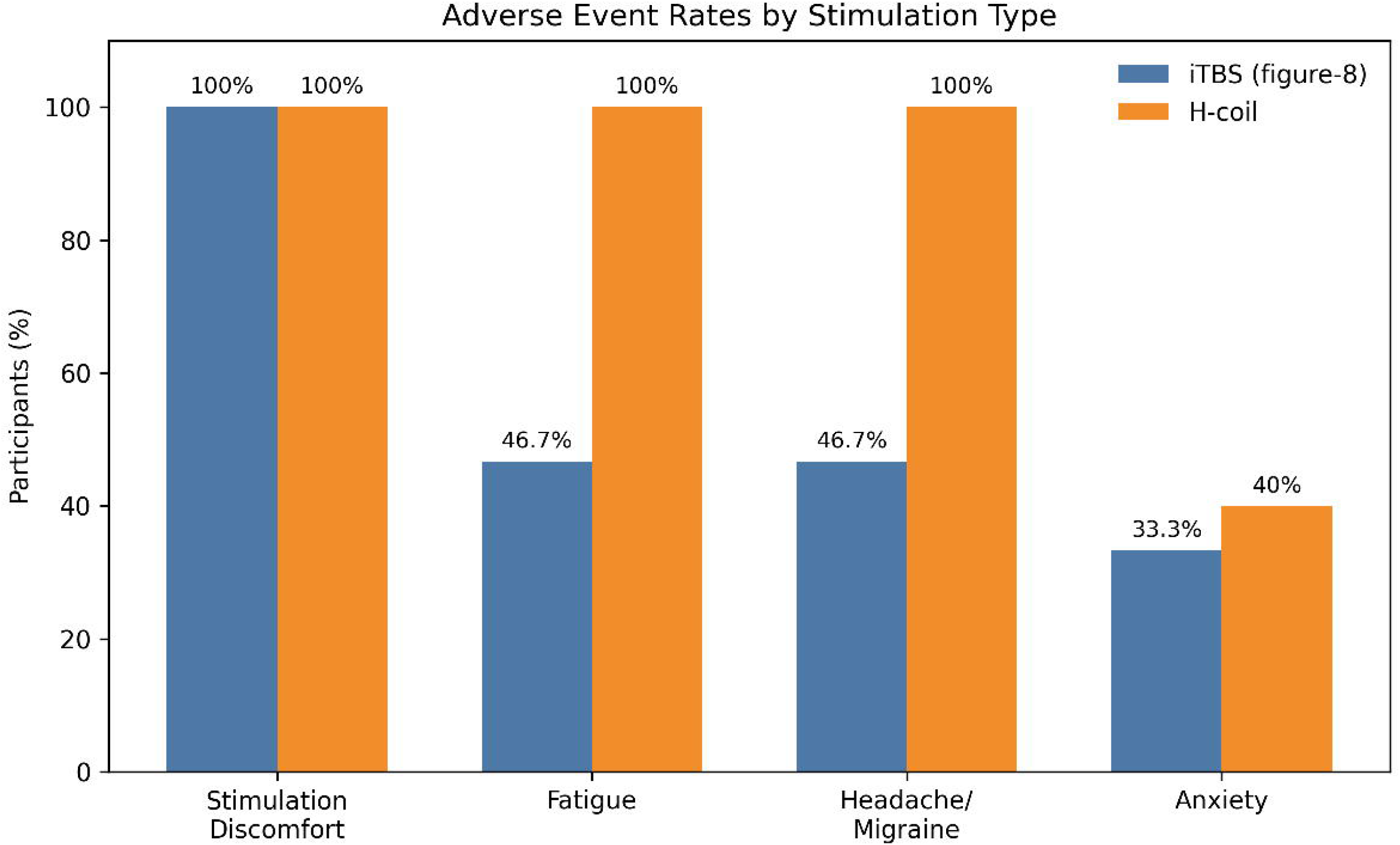

## 4. Discussion

This pilot randomized trial provides preliminary evidence that a head-to-head comparison of left-DLPFC iTBS and H7-coil rTMS is feasible in a treatment-resistant depression setting. Recruitment, randomization, adherence to the stimulation schedule, and retention across follow-up visits were all satisfactory, supporting the practicality of conducting a larger efficacy trial using this design.

Clinical outcomes should be interpreted as preliminary estimates rather than indicators of comparative efficacy. Although both groups showed symptomatic improvement over the six-week treatment period and through follow-up, the limited sample size does not allow conclusions about whether one modality performs better than the other. The absence of significant between-group differences is fully compatible with a true lack of difference, a modest difference undetectable with this sample size, or simply statistical underpowering. As such, these findings primarily inform the likely variability of outcome measures and the design parameters required for a fully powered trial. Exploratory cognitive analyses suggested that higher baseline executive functioning was associated with early remission, an effect that was primarily observed in the iTBS (figure-8 coil) group. Although these associations were not robust in adjusted models and must be interpreted cautiously given the small sample size, they raise the possibility that focal DLPFC iTBS may engage prefrontal cognitive control networks as part of its therapeutic mechanism. In contrast, the absence of similar cognitive predictors in the H-coil group suggests that medial prefrontal stimulation may exert antidepressant effects through alternative network pathways less directly dependent on baseline executive functioning. These preliminary signals should be viewed as hypothesis-generating and may help refine mechanistic hypotheses and cognitive targets for future adequately powered trials.

These findings extend prior work on non-invasive brain stimulation in major depression. The landmark THREE-D trial demonstrated non-inferiority of iTBS relative to 10 Hz rTMS, supporting iTBS as a time-efficient protocol in clinical practice [21]. In contrast, the present pilot study was designed to generate preliminary comparative estimates rather than to formally evaluate relative efficacy between modalities. While both groups showed symptomatic improvement over time, the sample size limits the inferences that can be drawn about potential differences, and the findings are best viewed as informing the parameters and design considerations of a future, fully powered trial. Evidence from multicenter studies of H1-coil rTMS has demonstrated efficacy over sham [9], and meta-analyses indicate broadly comparable outcomes between H1-coil rTMS and figure-8 rTMS, although effect sizes vary across trials [16]. Far fewer data are available specifically for the H7-coil; however, medial prefrontal targets such as the DMPFC have been examined in prior rTMS studies in TRD, supporting their clinical relevance, although direct comparisons with DLPFC stimulation remain scarce[22]. In this context, the present pilot trial provides early randomized observations within a single trial framework, informing the feasibility and design considerations of future adequately powered head-to-head studies targeting lateral versus medial prefrontal circuits.

Although exploratory, cognitive analyses indicated that remitters outperformed non-remitters on a composite executive index, with a trend toward better verbal fluency. These patterns, observed primarily in the iTBS rTMS group, align with prior studies linking executive functioning and frontostriatal integrity to treatment outcomes with both rTMS and pharmacotherapy. Experimental evidence further suggests that stimulation over the DLPFC can enhance working memory and modulate frontoparietal networks in healthy individuals [23]. Preserved executive function may therefore index network integrity and plasticity capacity, facilitating clinical response. Conversely, impaired executive functioning may represent entrenched network dysfunction, reducing treatment responsiveness. However, given the modest sample size and lack of robust statistical significance, these findings should be interpreted as hypothesis-generating.

Adverse events were frequently reported in both treatment arms, with some differences in their distribution across stimulation approaches. Stimulation-site discomfort was reported by all participants in both groups. Fatigue and headache or migraine were more commonly reported in the H-coil group than in the iTBS group (100% vs 46.7% for both outcomes). Mean pain ratings were comparable between groups (3.91 for iTBS and 3.57 for H-coil), suggesting similar levels of perceived stimulation discomfort. All adverse events were rated as mild to moderate in severity and were transient. No serious adverse events occurred, and no participants discontinued treatment due to adverse effects.

The present findings provide early descriptive information on symptom change with both iTBS and H7 rTMS in TRD. Although the study was not designed to determine relative clinical effectiveness, the response and remission rates observed here (40–50% and 20%, respectively) are broadly consistent with those reported in larger trials of rTMS for TRD [8]. These preliminary patterns may help contextualize expected variability and inform key design parameters for a future adequately powered comparative study. Considerations such as session duration, equipment availability, and tolerability remain practical factors for future trial planning, but the current results are not intended to guide clinical decision-making. Instead, they underscore the value of larger trials to clarify whether the two modalities differ meaningfully in their clinical outcomes.

iTBS and H7 rTMS differ markedly in terms of stimulation targets and field distribution. iTBS delivered over the left DLPFC primarily targets lateral prefrontal and frontoparietal networks, yet converging evidence shows that DLPFC stimulation can modulate cingulate circuitry indirectly through distributed network connectivity. In particular, the antidepressant efficacy of left-DLPFC TMS targets has been linked to their intrinsic functional connectivity (anticorrelation) with the subgenual cingulate, supporting a network mechanism rather than a strictly local one [24]. Related imaging and connectivity work also supports remote engagement of medial/cingulate regions following lateral prefrontal stimulation [25].

By contrast, H7-coil rTMS is intended to engage more medial prefrontal circuits (DMPFC) and potentially anterior cingulate regions, although this inference relies largely on electric-field modeling and known frontocingulate network architecture rather than direct evidence of focal cingulate stimulation [26]. Despite these anatomical and field-distribution distinctions, no clear differences in clinical outcomes emerged in this pilot sample. This may reflect convergent modulation of downstream networks implicated in depression (e.g., frontostriatal and frontocingulate circuits), such that different stimulation profiles can yield overlapping clinical effects at the network level [24].

Beyond coil design, the present trial can also be viewed in the context of prior work comparing lateral versus medial prefrontal stimulation targets in depression. Several studies have suggested that stimulation of the DMPFC may be particularly relevant for patients with prominent affective, self-referential, or cognitive control disturbances [22]. Connectivity-based targeting studies have shown that DMPFC stimulation engages frontocingulate networks implicated in mood regulation, while lateral DLPFC stimulation preferentially modulates frontoparietal control networks [24][27]. Across the existing literature, antidepressant effects reported with DMPFC-targeted stimulation appear to fall within a similar range to those observed with DLPFC-targeted rTMS, although direct head-to-head comparisons are scarce and substantial heterogeneity exists in patient characteristics, stimulation parameters, and targeting approaches. In this context, the present pilot trial adds preliminary randomized observations suggesting that both lateral and medial prefrontal stimulation strategies can be delivered within a single RCT framework, with acceptable feasibility and retention, while highlighting the need for adequately powered studies to determine whether specific patient subgroups may preferentially benefit from one target over the other.

This study has several limitations. The sample size was small and not powered to detect modest between-group differences, which limits the strength of the inferences that can be drawn from the absence of statistically significant effects. The single-site design limits generalizability, and blinding was not feasible due to differing coil configurations. Cognitive analyses were exploratory and should be replicated in larger trials. Future work may benefit from integrating neurophysiological or imaging biomarkers to better characterize treatment mechanisms and predictors of response. In particular, the role of cognitive functioning, as a trait predictor, treatment mediator, or therapeutic target, warrants further investigation.

## 5. Conclusions

This pilot randomized trial demonstrates that a head-to-head comparison of iTBS and H7-coil rTMS is feasible, with satisfactory recruitment, adherence, and retention across treatment and follow-up. Clinical outcomes should be interpreted as preliminary estimates rather than evidence of comparable efficacy, given the limited sample size. Notably, exploratory cognitive findings suggested that baseline executive functioning may relate to treatment response, particularly in the iTBS group. These patterns of symptom change and cognitive associations provide valuable information for planning a future adequately powered randomized trial. Larger studies will be required to determine whether the two modalities differ meaningfully in clinical outcome and to clarify the role of cognitive functioning in predicting treatment response.

## Data Availability

All data produced in the present study are available upon reasonable request to the authors

## Acknowledgements

The authors wish to sincerely thank the clinical nursing team of the neuromodulation unit for their dedication and professionalism in delivering the treatments throughout the study. We are especially grateful to Ana Baker, who played a central role in coordinating the project at the clinical level and ensuring the smooth implementation of the protocol.

## Author Contributions

JPM conceptualized the study. EB, VDJ and JPM contributed to data collection, supervision, or analysis. VDJ, EB, and JPM drafted the manuscript. FT-D, DJA, FL, EBA, DKN, DB, TB, MA, ZJD, and PL contributed to interpretation of the results and critical revision of the manuscript. All authors reviewed and approved the final version.

